# Protocol on an Integrative review on nomenclature and outcomes in children with complex critical illness in Paediatric Intensive Care - The basis for consensus definition

**DOI:** 10.1101/2024.04.29.24306579

**Authors:** Sofia Cuevas-Asturias, Will Tremlett, Hannah K Mitchell, Claire Rafferty, Padmanabhan Ramnarayan, Natalie Pattison

**Affiliations:** Imperial College London; Birmingham Children’s hospital; Institute of Child Health, University College London, London, UK; The Bristol Royal Hospital for Children

**Keywords:** Complex critical illness, Paediatric intensive care, children with medical complexity, severe chronic illness, PICU

## Abstract

Paediatric Critical Care (PCC) supports the recovery of children with severe illness. Nationally, there are 30 PCC units with a total of approximately four hundred beds. There is constant demand for these beds with a mean five-day length of stay and admissions increasing at a greater rate than age-specific population growth [1, 2]. Prolonged stay patients account for approximately half of all PCC patient bed days [3].

Children with complex critical illness (CCI) need input from multiple different teams alongside support for their family [4, 5]. CCI often become prolonged PCC-stay patients too [6]. Internationally, there is variation in the definition of CCI [4, 8], this creates service variation and tensions around what resources can be provided including discharge planning, provision, and support.

**Objective:** The face of Paediatric Critical Care, in the UK and internationally has changed over the course of the last ten years with a growing cohort of complex critically ill patients. This integrative review aims to look at current nomenclature, definitions, and outcome measures of priority in this undefined patient population.

**Inclusion criteria:** All types of studies looking at children with complex critical illness (age <18 years) admitted to any paediatric intensive care unit (PICU).

**Methods:** The review is registered on Prospero. Medline, Embase, Maternity and Infant care, The Cochrane library, the Cumulative Index to Nursing and Allied Health literature (CINAHL) and Trip database will be searched from 2014 to May 2024.

Search limits will include all languages, exclude the setting of neonatal intensive care and age>18 years old. Four independent reviewers will screen citations for eligible studies and perform data extraction. The final search strategy will be developed in Medline and peer-reviewed by a health research librarian not involved in the study. This will be translated to other databases as appropriate.

**Author approval:** All authors have seen and approved the manuscript.

## Strengths and limitations of this study

1. An integrative review methodology has been used to develop a comprehensive understanding of the literature which will be used to develop further work in this area.
2. Using a rigorous and stepwise approach, the whole spectrum of scientific publications on complex critical patients in paediatric intensive care will be reviewed, ensuring this study is as comprehensive as possible. This includes quantitative, qualitative, theoretical, and grey literature.
3. A limitation of this integrative review is the use of many terms to describe complex critical patients in the literature resulting in a high number of publications on this topic.

## Introduction

Paediatric Critical Care (PCC) has 30 units throughout the UK, with a total of approximately 400 beds [2]. The overall occupancy of PCC beds runs critically high (above 85% occupancy) most of the year and studies have shown an increasing demand for PCC beds with concern that demand may soon outstrip resources [2]. One reason for increasing demand is greater numbers of a heterogenous group of children with complex critical illness (CCI) who often have prolonged lengths of stay. Existing classifications systems of CCI patients are variable and do not allow for accurate documentation of incidence of PICU admission and subsequent mortality or morbidity [9].

Definition for a prolonged length of stay (PLOS) PCC admission varies from >14 to >28 days. Over the last two decades, PLOS PICU admissions have increased significantly and now account for between 42-51% of PICU patient-days [3]. PLOS PICU patients have a high resource utilisation and a median overall hospital length of stay of 98 days[3]. PCC has a decreasing overall trend in mortality, but PLOS patients have significantly higher mortality than the general PICU population [3, 10].

### Children with complex critical illness (CCI)

Children with CCI present unique challenges to PCC. These challenges include:

- (Frequently) prolonged length of stay
- Complex medical regimens
- Complicated family dynamics
- Multiple specialist and allied healthcare professional (AHP) input amongst others[4].

A national survey published in 2024 with the Paediatric Critical Care Society Study Group (PCCS-SG) looked at provision for children with CCI. This showed variable patient identification, management, and care pathways[4]. Currently, there are two PCC units in the UK with a specific multidisciplinary toolkit for the management of children with CCI[4]. Due to a lack of definition for children with CCI, the economic impact, mortality, or morbidity of this patient group is unknown.

This integrative review will provide a detailed analysis of evidence around nomenclature, definitions of this patient cohort alongside outcomes of measurement that have been used in the literature. This will aid the development of a definition via a consensus group.

A preliminary search of MEDLINE, Cochrane database of systematic reviews, COMET initiative, Prospero and JBI Evidence Synthesis was conducted and no current or underway systematic reviews or integrative reviews on the topic were identified. Zorko et al conducted a scoping review in January 2023 which looked at Chronic critical illness within PICU[11], this differs as it does not encompass medical complexity at admission or repeated admissions.

The objective of this integrative review is to assess the extent of the literature within paediatric complex critical illness and associated outcomes of priority to aid the development of a definition and core outcomes set for this patient group.

## Research question

This integrative review will aim to answer the following questions:

1. How are children with complex critical illness defined in current literature? As there is no current standard of this definition, this review will evaluate how medical complexity, chronic critical illness and prolonged PICU admissions have been defined.
2. What are the demographics and clinical characteristics of children with complex critical illness in PICU based on existing definitions?
3. Health outcomes of interest – what are the outcomes studied currently in this patient group?

The purpose of the integrative study is to use this knowledge to develop a pragmatic consensus definition. With this definition, a core outcomes data set will be developed for the UK.

## Eligibility criteria

### Population

Studies looking at critically ill children (age<18 years) admitted to any paediatric intensive care unit (PICU) identified with the following terms:

- Paediatric complex critical illness
- Complex chronic conditions
- Prolonged or long-stay PICU admission
- Medical complexity
- Severe or chronic critical illness
- Severe neurologic impairment
- Technology dependent children

These have been adapted from Edwards 2022 paper as per Table 1 below.

**Table 1:** Common Classifications of Children with Severe Chronic Illness and Their Illnesses (adapted from Edwards et al. PCCM 2022)[9].

| Classification | Commonly Accepted Definition |
| --- | --- |
| **Complex chronic condition** | Any medical condition that can be expected to last at least 12 months (unless death intervenes) and to involve either several different organ systems or one organ system severely enough to require specialty paediatric care and probably some period of hospitalisation in a tertiary care centre[12] |
| **Children with medical complexity** | Those with 1) chronic conditions (specifically of the more severe or CCC type); 2) substantial |
|  | functional limitations; 3) increased health and other service needs; and 4) increased healthcare costs[13] |
| **Chronic critical illness** | This term was taken from adult medicine which was developed in 1985[14] and has a variable definition in paediatric literature[11]. Current definitions in literature include:<br>Conditions in which children have both of the following:<br>- Hospitalised > 28 days in a neonatal ICU or > 14 in a PCC or who have had a prolonged ICU stay (not defined) and two or more subsequent admissions (PCC or general ward)<br>- Ongoing dependence on technologies to sustain vital functions[15]. |
| **Severe neurologic impairment** | Another definition with wide variation. One definition:<br>CNS disorders that arise in childhood resulting in motor and cognitive impairment and medical complexity, where much assistance is required with activities of daily living. The impairment is permanent and can be progressive or static[16, 17]. |
| **Technology dependent children** | Another definition with wide variation. One definition:<br>Technology dependency refers to a wide range of clinical technology to support biological functioning across a dependency continuum, for a range of clinical conditions. It is commonly initiated within a complex biopsychosocial context and has wide-ranging sequelae for the child and family, and health and social care delivery.[18] |

#### Intervention, Comparator, Outcome

Any or none. Outcomes will be identified using the COMET theoretical framework to guide the development of a core outcomes set alongside definition development.

##### Concept

The concept is the intersection between acute and chronic illness in this population and their associated outcome measures.

##### Context

Studies within paediatric intensive care. We will be conducting an international integrative review looking at PICU CCI worldwide. However, this will be specifically for the development of a definition within the UK.

###### Exclusions

The study evaluates only adult population or evaluates adults and paediatric populations but does not report separate data for each population.

### Types of Sources

This integrative review will consider experimental studies, randomised controlled trials, non-randomised controlled trials, before and after studies and interrupted time-series studies. In addition, analytical observational studies including prospective and retrospective cohort studies, case-control studies and analytical cross-sectional studies will be considered for inclusion. This review will also consider descriptive observational study designs including case series, individual case reports and descriptive cross-sectional studies for inclusion. Qualitative studies will also be considered that focus on qualitative data including, but not limited to, designs such as grounded theory, ethnography, qualitative description, and action research.

In addition, systematic reviews that meet the inclusion criteria will also be considered, depending on the research question. Text and opinion papers will also be considered for inclusion in this integrative review.

## Methods and analysis

The protocol was developed using integrative review published methodology[19-23]. An accurate audit trail will be kept allowing for reproducibility[22]. The integrative review will be registered on Prospero. The proposed integrative review will be reported with guidance from the standards of the Preferred Reporting Items for Systematic Review and Meta-analysis (PRISMA) Statement.

### Search strategy

The search strategy will aim to locate both published and unpublished studies. An initial limited search of Ovid MEDLINE will be done to identify articles on the topic. The text words contained in the titles and abstracts of relevant articles, and the index terms used to describe the articles were used to develop a full search strategy for The Cochrane library, the Cumulative Index to Nursing and Allied Health literature (CINAHL), MEDLINE, EMBASE, Trip database (see Appendix 1).

The search strategy, including all identified keywords and index terms, will be adapted for each included database and/or information source. The reference list of all included sources of evidence will be screened for additional studies.

Ongoing consultation with a health librarian will aid specificity and comprehensiveness of the search. Each search strategy will be within the Prospero registration appendix to allow another reviewer the ability to replicate and/or evaluate the search. Studies published since 2014 will be included as complex critical illness is a novel concept within paediatric critical care.

#### Study/Source of Evidence selection

Following the search, all identified citations will be collated and uploaded into Covidence and duplicates removed. Following a pilot test, titles and abstracts will be screened by three independent reviewers for assessment against the inclusion criteria. All sampling decisions will be transparent and justified with a PRISMA search flow diagram[23].

The full text of selected citations will be assessed in detail against the inclusion criteria by two or more independent reviewers. Reasons for exclusion of sources of evidence at full text review that do not meet the inclusion criteria will be recorded and reported in the integrative review. The search results and study inclusion process will be reported in full in the final integrative review and presented in a Preferred Reporting Items for Systematic Reviews and Meta-analyses (PRISMA) flow diagram[7].

### Data Extraction

Data will be extracted from papers included in the integrative review by three independent reviewers using the systematic review software tool Covidence. The data extracted will include specific details about the participants, concept, context, study methods and key findings relevant to this integrative review.

A draft extraction form is provided (See Appendix 3). The draft data extraction tool will be modified as required during each data extraction process. Modifications will be detailed in the integrative review. Any disagreements that arise between the reviewers will be resolved through discussion, or with an additional reviewer/s. If appropriate, authors of papers will be contacted to request missing or additional data, where required. Critical appraisal of individual sources of evidence will be done.

#### Quality assessment

Eight studies (four quantitative and four qualitative studies) will be randomly picked to refine the CASP scoring criteria by all four reviewers. Consistencies and inconsistencies between reviewers will be noted, alongside this, the scoring system will be modified according to problems encountered.

Data will also be clustered geographically into related subgroups to identify themes whilst maintaining a detailed audit trail. A narrative summary will accompany the tabulated results and will describe how the results relate to the reviews objectives and questions. We will consider conducting a sensitivity analysis for qualitative studies.

### Data synthesis

The quality and validity of studies selected will be assessed using the critical appraisal skills programme checklists (CASP) to mitigate bias. The evidence will be presented in a review table with quality scores for each source incorporated into the data table under a column heading that identifies the method of evaluation used. All reviewers will be involved in the quality evaluation.

Quantitative data will be pooled statistically if there is sufficient available data for meta-analysis. If statistical pooling is not possible, the findings will be presented in narrative form. Data will also be clustered geographically into related subgroups to identify themes whilst maintaining a detailed audit trail. A narrative summary will accompany the tabulated results and will describe how the results relate to the reviews objectives and questions.

Analysis of the findings will include identifying strengths and weaknesses of the current literature regarding defining complex critical patients and core outcomes. Synthesis of the findings will provide new understandings of this topic to form a basis for consensus definition development and core outcomes set [15].

### Outcomes

The main outcome of the research will provide an evidence base to inform and influence development of a definition for paediatric complex critical illness. As there is no current standard of this definition, this review will evaluate how medical complexity, chronic critical illness and prolonged PICU admissions have been defined. We will also look at how the definition was developed and/or validated by primary study.

Following this, the findings of this review will be used to inform a future programme of research aimed at improving the identification, management, and outcomes of paediatric complex critical illness patients.

Additional outcomes will be any objective measure of outcomes including:

- Health related outcomes using validated tools where possible (e.g. functional status, severity of illness, co-morbidities, quality of life, symptom burden, unmet needs, satisfaction, rates of hospitalisation)
- Process outcomes (e.g. patient behavioural uptake, quality of care, training, and education; costs and resource utilisation).

Interventions will be pooled to look at common attributes and potential benefits/shortfalls to aid in development of core outcomes set for this population group.

The new knowledge produced may contribute to education and training of undergraduate and postgraduate paediatric intensive care multidisciplinary health professionals and allied health and social care professionals.

We aim to publish the protocol and integrative review findings in academic paediatric critical care literature to ensure that findings are internationally available to practitioners and researchers.

### Patient and Public Involvement

This protocol was developed with public and patient involvement. They were involved from first principal onwards. The research questions were informed from previous patient engagement in PICU. Parents of children with CCI have commented on the methods and have been integral to protocol finalisation. They will continue to be involved in future work using the integrative review for development of a definition for complex critical patients in PICU. They will also aid in dissemination through patient and public networks within complex critical patients.

## Discussion and Conclusion

The discussion will include comparisons, and contrasts of the findings of the review with background literature. Recommendations and implications for future research practice, core outcomes set and consensus definition working group development will be made.

### Ethics and dissemination

This proposal seeks to conduct an integrative review that will not involve human or animal subjects. Therefore, review by an ethics committee is not required. We aim to publish the results in peer-reviewed journals and presentations at conferences in the United Kingdom and internationally.

## Supporting information

Appendix with search strategy and data extration form

## Data Availability

All data produced in the present study are available upon reasonable request to the authors

## Ethics statements

Patient consent for publication is not applicable.

## Contributors

SCA drafted the overall research idea and wrote the initial draft of the protocol. PR and NP contributed content and edited the content. All authors revised and approved the final manuscript.

