## Appendix with search strategy and data extration form for "Protocol on an Integrative review on nomenclature and outcomes in children with complex critical illness in Paediatric Intensive Care - The basis for consensus definition"

### Appendix 1: Search Strategy

#### For Medline:

1. exp Critical Care/
2. Critical* Care.mp.
3. critical* ill*.mp.
4. ICU.mp.
5. intensive care units/
6. exp Intensive Care Units, Pediatric/
7. PICU.mp.    7417
8. PCCU.mp.    49
9. ((p?ediatric or child*) and (intensive or ICU)).mp.
10. 1 or 2 or 3 or 4 or 5 or 6 or 7 or 8 or 9
11. exp adolescent/ or exp child/ or exp infan*/
12. (child* or teen* or infan* or adolescen* or p?ediatric or young or youth or Juvenile).mp.
13. 11 or 12
14. exp Chronic Disease/
15. chronic disease*.mp.
16. (complex adj3 need*).mp.
17. (complex adj2 condition*).mp.
18. (complex adj3 condition*).mp.
19. Life-limit*.mp.
20. Lifelimit*.mp.
21. (chronic adj2 illness*).mp.
22. Chronic health.mp.
23. medical complexity.mp.
24. patient complexity.mp.
25. (multi adj3 morbid*).mp
26. Long-term illness.mp.
27. Technology-Dependent Child*.mp.
28. medically fragile.mp.
29. (severe adj2 condition*).mp.
30. neurodisability.mp.
31. prolonged-stay.mp.
32. 14 or 15 or 16 or 17 or 18 or 19 or 20 or 21 or 22 or 23 or 24 or 25 or 26 or 27 or 28 or 29 or 30 or 31
33. 10 and 13 and 32
34. limit 33 to yr="2014 -Current"

### Appendix 2: Draft data extraction form

| **Source** | **Eligibility** | **Study Characteristics** | **Methods** | **Patient Demographics** | **Population definition & Key findings or outcome of interest** | **Miscellaneous** |
| --- | --- | --- | --- | --- | --- | --- |
| Study ID (created by review author). | Confirm eligibility for review. | Author name | Study design. | Total number and groups if applicable | How is the study population defined. | Funding source. |
| Review author ID (created by review author). | Reason for exclusion | Title | Total study duration. | Reason for PICU admission | Definition of paediatric complex critical illness | Key conclusions of the study authors. |
| Citation and contact details. |  | Country of origin | Sequence generation. | Diagnostic criteria. | Definition of prolonged PICU admission | Miscellaneous comments from the study authors. |
|  |  | Journal and year of publication | Allocation sequence concealment. | Age. | Definition of medical complexity in PICU | References to other relevant studies. |
|  |  | Clinical setting/type of PICU | Blinding. | Sex. | How the definition was developed and/or validated by primary study. | Correspondence required. |
|  |  |  | Other concerns about bias*. | Functional status (using validated tools such as functional status score) | *For each outcome of interest*:  Outcome definition (with diagnostic criteria if relevant). | Miscellaneous comments by the review authors. |
|  |  |  |  | Severity of illness (using validated tools) | Unit of measurement (if relevant). |  |
|  |  |  |  | Co-morbidity |  |  |
